# Lymphatic Spread Patterns in HPV+ vs. HPV− Oropharyngeal Squamous Cell Carcinoma: A Multi-Center European Analysis

**DOI:** 10.64898/2026.09.29.26364302

**Authors:** Yoel Pérez Haas, Roman Ludwig, Esmée L. Looman, Panagiotis Balermpas, Sergi Benavente, Adrian Schubert, Dorothea Barbatei, Laurence Bauwens, Olgun Elicin, Matthias S. Dettmer, Roland Giger, Vincent Grégoire, Jan Unkelbach

## Abstract

**Background:** Human papillomavirus (HPV)–positive oropharyngeal squamous cell carcinoma (OPSCC) shows higher prevalence of lymph node metastases than HPV-negative OPSCC. It remains unclear whether HPV-positive and HPV-negative OPSCC differ in occult metastasis risk, which is relevant to personalize elective nodal treatment volumes.

**Methods:** We performed a retrospective analysis of 780 untreated OPSCC patients from four European centers. Patterns of lymph node level involvement (LNLs I, II, III, IV, V, VII) were assessed using a consensus diagnosis integrating imaging (CT, MRI, PET) and pathological findings from surgically treated patients. Occult metastases were assessed in 288 surgical patients with paired clinical and pathological information.

**Results:** HPV-positive patients more frequently presented with ipsilateral LNL II involvement, particularly in early T-stage disease (80.6% [75.6–84.8] vs. 54.8% [47.2–62.2]). However, the probability of downstream involvement in LNLs III and IV conditioned on upstream LNL involvement was similar between HPV-positive and HPV-negative patients. Similarly, contralateral involvement conditioned on midline extension of the primary tumor was similar. The overall proportion of occult metastases, defined as clinically false-negative LNLs divided by pathologically positive LNLs, was lower in HPV-positive patients (24.7% [19.7–30.1] vs. 48.7% [39.7–57.8]).

**Conclusions:** HPV-positive OPSCC showed higher prevalence of LNL involvement, especially in ipsilateral LNL II and early T-stage tumors. The data to directly compare occult metastasis risk has important limitations and does not allow definitive conclusions. However, the presented data suggests that higher prevalence likely does not translate into higher risk of occult metastases for the most common clinical diagnoses and therefore does not support larger elective nodal treatment volumes compared with HPV-negative tumors.

## 1 Introduction

Head and neck squamous cell carcinomas (HNSCC) frequently spread through the lymphatic system, leading to metastatic involvement of cervical lymph node levels (LNLs) [1, 2, 3]. In oropharyngeal squamous cell carcinoma (OPSCC), lymph node metastases are present in approximately 70% of patients [4, 5, 6]. In addition to clinically detected metastases, treatment includes LNLs at risk of harboring occult metastases, which is referred to as prophylactic neck dissection or elective nodal irradiation (ENI).

Whereas current guidelines on ENI recommend large elective nodal clinical target volumes (CTV-N), volume-deescalation strategies and personalization of the elective CTV-N have recently gained attention. In addition to studies on unilateral versus bilateral ENI, this includes the EVADER trial [7] and the DeEscO trial [8], which personalize the CTV-N based on the clinical lymph node involvement as well as lateralization, T-category, and/or subsite of the primary tumor. In the context of personalized CTV-N definition, an open question is whether the risk of occult metastases and the appropriate CTV-N differs between HPV-positive and HPV-negative OPSCC.

Human papillomavirus (HPV) is a major etiologic factor in OPSCC [9] and has reshaped the epidemiology of the disease over the past two decades [10, 11]. In addition to differences in patient characteristics and tumor biology [12, 13, 14], HPV-positive OPSCC is associated with markedly improved treatment response and survival [9, 15, 16]. These distinctions support the notion that HPV-positive and HPV-negative OPSCC represent biologically and clinically distinct entities.

HPV-positive OPSCC more often presents with earlier T-category primaries but more extensive nodal disease compared to HPV-negative tumors [9, 17, 18]. However, the implications of this observation for ENI are unclear. Higher prevalence of nodal disease does not per se imply higher probability of occult metastases in a clinically negative LNL for an individual patient. Current international guidelines recommend the same CTV-N irrespective of HPV-status [19, 20].

Although several studies have investigated occult metastasis rates in HPV-positive patients [21, 22, 23], personalized quantification of occult metastasis risk, taking into account HPV but also other known risk factors, is inherently limited as it would require a very large cohort of surgically treated patients. In this work, we present a detailed multi-institutional retrospective analysis comparing LNL involvement between HPV-positive and HPV-negative OPSCC in 780 OPSCC patients. In the discussion section, we discuss the relation between observed patterns of LNL involvement and the risk of occult metastases, and the implications of the findings for personalized CTV-N definition.

## 2 Methods and Materials

### 2.1 Patient cohort

We performed a retrospective, multi-institutional analysis of patient-level data from four European centers. Eligible patients had newly diagnosed, untreated OPSCC (ICD codes C01, C09, C10). For each patient, involvement was recorded for LNLs I, II, III, IV, V, and VII. HPV status was determined with p16 immunohistochemistry as a surrogate. Throughout this manuscript, p16-positive is referred to as HPV+ and p16-negative as HPV–.

Data were contributed by the following participating centers:

1. **Centre Léon Bérard (CLB), Lyon, France**: 315 patients in total [24]. For 237 patients [17] both clinical involvement and pathological diagnoses after neck dissection for resected LNLs were available. For the remaining 78 patients, only pathological information was available and the lateralization of the primary tumor was unknown.
2. **Inselspital Bern (ISB), Bern, Switzerland**: 74 patients, with reported pathological and clinical involvement. However, in some cases multiple levels were resected en bloc and positive lymph nodes could not be uniquely assigned to a LNL [24].
3. **Vall d’Hebron Hospital (HVH), Barcelona, Spain**: 114 patients with clinical involvement reported, and pathological confirmation in a subset of 4 patients. [25].
4. **University Hospital Zurich (USZ), Zurich, Switzerland**: 277 patients with clinical involvement reported, and pathological confirmation in a subset of 2 patients. [6].

In total, 780 patients were included. The aggregated dataset is publicly accessible through the LyProX platform (https://lyprox.org/). Details of the datasets are described in the respective publications.

For analysis, T-category was grouped into early (T1–T2) and late (T3–T4) stages, which are defined consistently across TNM editions.

### 2.2 Assessment of LNL Involvement

LNL involvement was assessed at the level of individual LNLs using three diagnostic methods:

- **Pathological assessment** after neck dissection. These data were available only for surgically treated patients and only for dissected LNLs.
- **Clinical involvement** based on computed tomography (CT), magnetic resonance imaging (MRI), positron emission tomography (PET), and fine needle aspiration (FNA). For datasets reporting involvement separately for multiple modalities, the clinical information was combined into a clinical consensus as described in Ludwig et al. [26].
- **Diagnostic consensus**: a per-LNL consensus diagnosis combining clinical involvement with pathological information when available. Pathological results were considered the gold standard and overwrote potentially conflicting information on clinical involvement.

### 2.3 Analysis of LNL Involvement

We performed a detailed analysis of the prevalence of LNL involvement depending on HPV status, Tstage, and lateralization of the primary tumor. In addition, correlations between LNLs were analyzed, i.e. probability of involvement of downstream LNLs depending on the involvement of upstream LNLs. These analyses were performed based on the diagnostic consensus including all patients.

Occult metastases were analyzed in the subset of patients and LNLs for which both clinical involvement and pathological assessment were available. Two quantities were considered:

- **Proportion of occult metastases among pathologically positive LNLs**: This quantity describes how often pathological involvement was missed clinically. It was calculated as the fraction of pathologically positive LNLs that were clinically negative, corresponding to one minus the sensitivity of the clinical diagnostic procedure. The analysis included all ipsilateral LNLs for which both clinical and pathological assessment were available.
- **Occult metastasis risk in clinically negative LNLs**: This quantity estimates the risk that a clinically negative LNL harbors pathological disease. It was calculated as the number of clinically negative but pathologically positive LNLs divided by the number of patients with a given clinical diagnosis (in whom the LNL of interest is clinically negative). This analysis was restricted to 288 patients for whom both clinical and pathological diagnoses were available for ipsilateral LNLs II, III, and IV. Contralateral occult metastases were not analyzed due to the small number of patients with bilateral neck dissection.

### 2.4 Statistical Analysis

Given the retrospective nature of the study, the analysis was primarily descriptive. LNL involvement was reported as counts and percentages in patient subgroups, with statistical uncertainties reported as 95% Wilson confidence intervals.

## 3 Results

Of the 780 included patients, 456 (58%) were HPV+ and 324 (42%) were HPV– (Table 1, Figure 1). Early T-stages (T1–T2) were more common among HPV+ patients (284/456, 62.3%) than among HPV– patients (166/324, 51.2%).

**Table 1:** Ipsilateral LNL involvement based on diagnostic consensus, stratified by T-stage and HPV status. For each subgroup and LNL, the first number is the count of involved patients and the second is the proportion within the subgroup. For LNLs I–V, percentages are calculated using the row-specific patient number *n*. For LNL VII, only 629 ipsilateral diagnoses were available; percentages are therefore calculated using subgroup-specific denominators: All, 629; T1/T2, 338; T3/T4, 291; HPV+, 353; HPV–, 276; T1/T2 HPV+, 207; T1/T2 HPV–, 131; T3/T4 HPV+, 146; T3/T4 HPV–, 145.

|  | n | I |  | II |  | III |  | IV |  | V |  | VII |  |
| --- | --- | --- | --- | --- | --- | --- | --- | --- | --- | --- | --- | --- | --- |
| All | 780 | 76 | 9.7% | 575 | 73.7% | 254 | 32.6% | 74 | 9.5% | 61 | 7.8% | 52 | 8.3% |
| T1/T2 | 450 | 21 | 4.7% | 320 | 71.1% | 123 | 27.3% | 35 | 7.8% | 29 | 6.4% | 20 | 5.9% |
| T3/T4 | 330 | 55 | 16.7% | 255 | 77.3% | 131 | 39.7% | 39 | 11.8% | 32 | 9.7% | 32 | 11.0% |
| HPV+ | 456 | 37 | 8.1% | 372 | 81.6% | 154 | 33.8% | 42 | 9.2% | 40 | 8.8% | 27 | 7.6% |
| HPV- | 324 | 39 | 12.0% | 203 | 62.7% | 100 | 30.9% | 32 | 9.9% | 21 | 6.5% | 25 | 9.1% |
| T1/T2 HPV+ | 284 | 12 | 4.2% | 229 | 80.6% | 86 | 30.3% | 21 | 7.4% | 22 | 7.7% | 13 | 6.3% |
| T1/T2 HPV- | 166 | 9 | 5.4% | 91 | 54.8% | 37 | 22.3% | 14 | 8.4% | 7 | 4.2% | 7 | 5.3% |
| T3/T4 HPV+ | 172 | 25 | 14.5% | 143 | 83.1% | 68 | 39.5% | 21 | 12.2% | 18 | 10.5% | 14 | 9.6% |
| T3/T4 HPV- | 158 | 30 | 19.0% | 112 | 70.9% | 63 | 39.9% | 18 | 11.4% | 14 | 8.9% | 18 | 12.4% |

**Figure 1:**
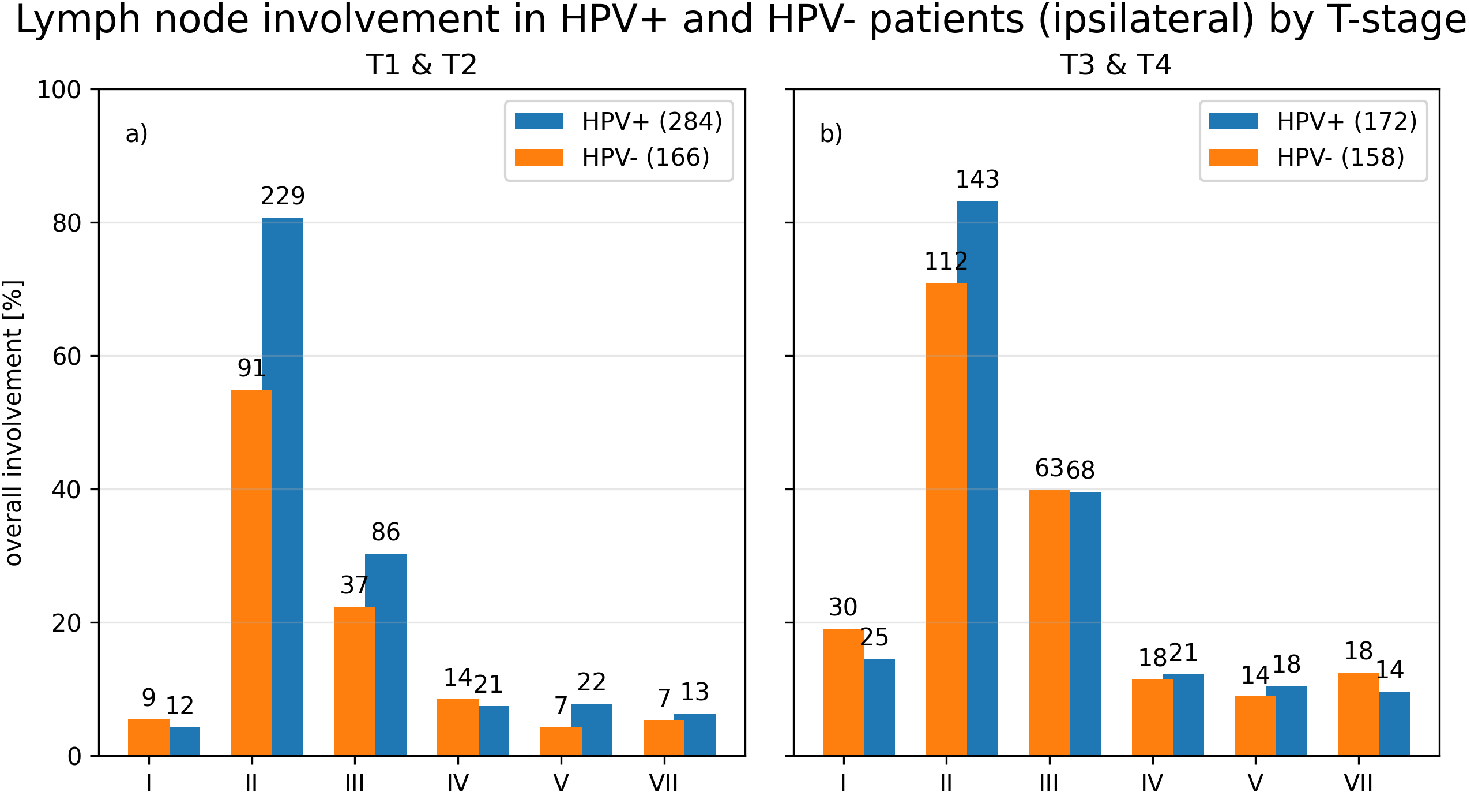
Ipsilateral involvement by LNL based on the diagnostic consensus, stratified by T-stage (early: T1–T2; late: T3–T4) and HPV status. Bars display proportions with absolute counts shown above each bar. Color scheme: HPV+ (blue), HPV– (orange).

### 3.1 Ipsilateral Involvement

Across all T-stages, ipsilateral LNL II involvement was higher in HPV+ than in HPV– patients (372/456, 81.6% vs. 203/324, 62.7%). This difference was most pronounced in early T-stages: HPV+ 229/284 (80.6% [75.6–84.8]) vs. HPV– 91/166 (54.8% [47.2–62.2]). In late T-stages, the difference was smaller: HPV+ 143/172 (83.1%, 95% CI: 76.8–88.0%) vs. HPV– 112/158 (70.9%, 95% CI: 63.4–77.4%).

For LNL III, a modest difference was observed in early T-stages (HPV+ 86/284, 30.2% vs. HPV– 37/166, 22.3%), which disappeared in late stages (HPV+ 68/172, 39.5% vs. HPV– 63/158, 39.9%). For LNLs I, IV, V, and VII, no clear differences in involvement by HPV status were observed after stratification by T-stage.

### 3.2 Contralateral Involvement

Figure 2 shows contralateral LNL involvement stratified by midline extension and HPV status. Midline extension was defined as primary tumor extension across the mid-sagittal plane. Only patients with known midline extension status were included in this analysis; 78 patients were excluded due to missing midline extension information. After stratification by midline extension, contralateral involvement patterns were similar between HPV+ and HPV– patients, with no evidence of increased contralateral spread associated with positive HPV status.

**Figure 2:**
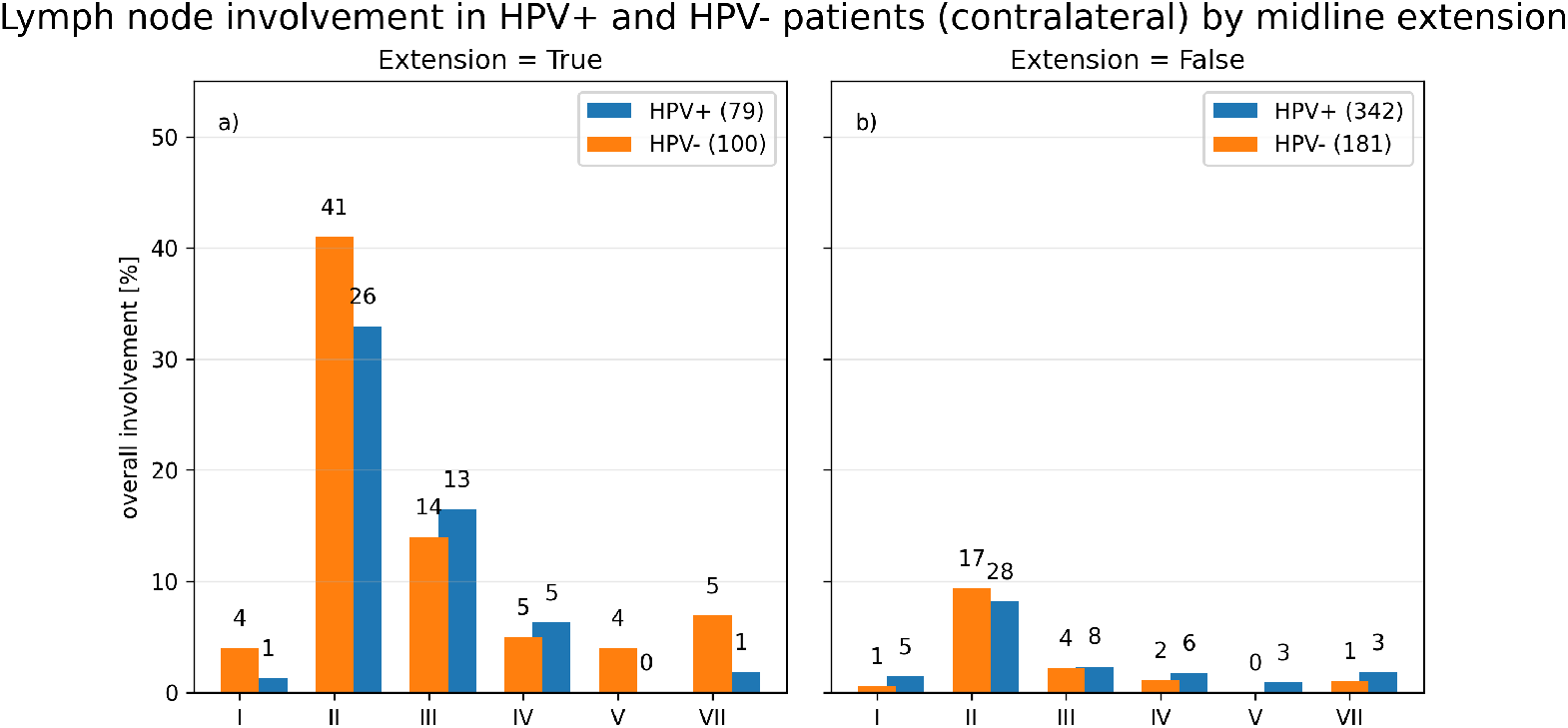
Contralateral LNL involvement based on the diagnostic consensus, stratified by midline extension and HPV status. Bars display proportions with absolute counts indicated above each bar. Color scheme: HPV+ (blue), HPV– (orange).

### 3.3 Downstream Involvement and Skip Metastases

Probabilities of downstream involvement and skip metastases are shown in Figure 3. Panels 3a and 3c display downstream involvement of LNL III given LNL II involved, and LNL IV given LNL III involved, respectively, whereas Panels 3b and 3d display skip metastasis probabilities in LNL III despite LNL II being uninvolved, and LNL IV despite LNL III being uninvolved, respectively. For all four cases, differences between HPV+ and HPV– patients were minimal.

**Figure 3:**
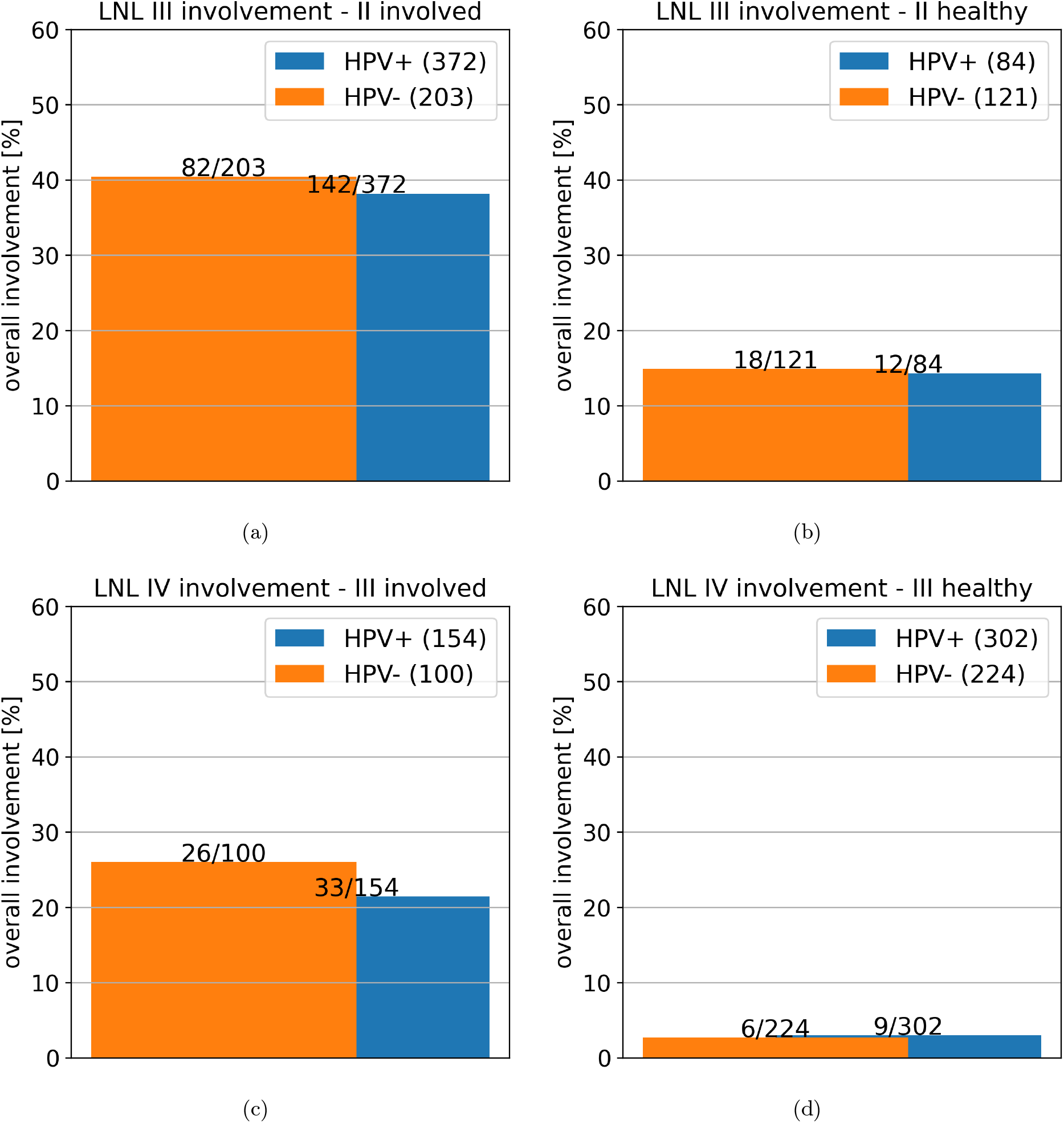
Downstream involvement and skip metastases by HPV status. Bars compare HPV+ (blue) and HPV– (orange). Downstream denotes involvement of a lower LNL given the immediately upstream LNL is involved; skip denotes involvement of a lower LNL despite the upstream LNL being uninvolved.

For LNL III involvement with LNL II involvement, the probabilities were 40.4% [33.9–47.3] for HPV– and 38.2% [33.4–43.2] for HPV+ patients. For LNL III involvement despite LNL II being uninvolved, the probabilities were 14.9% [9.6–22.3] and 14.3% [8.4–23.3], respectively. For LNL IV involvement given LNL III involvement, the probabilities were 26.0% [18.4–35.4] for HPV– and 21.4% [15.7–28.6] for HPV+ patients. Finally, for LNL IV involvement despite LNL III being uninvolved, the probabilities were 2.7% [1.2–5.7] and 3.0% [1.6–5.6], respectively.

### 3.4 Sensitivity of clinically detecting metastases

Table 2 summarizes the proportion of pathologically positive LNLs that were clinically negative, i.e. the occult metastasis rate among pathologically positive LNLs stratified by HPV status. Overall, a lower proportion of LNLs had occult involvement in HPV+ patients than in HPV– patients (24.7% [19.7–30.1] vs. 48.7% [39.7–57.8]).

**Table 2:**
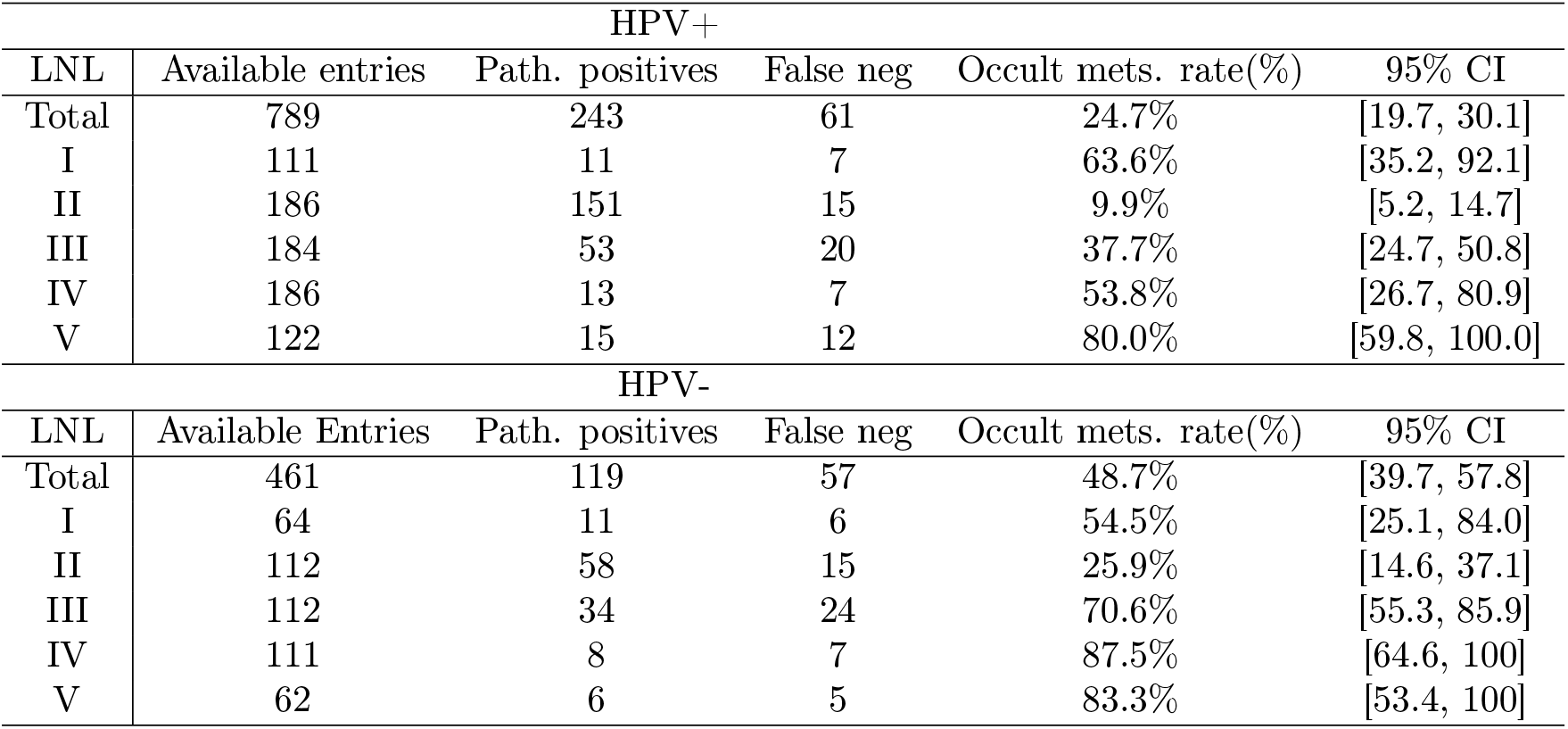
Sensitivity of clinically detecting metastases, reported as the proportion of occult metastases (false-negative rates) in HPV+ and HPV– patients by LNL. “Pathologically positives” indicates the total number of LNLs with a positive pathological diagnosis. “false-negatives” denotes clinically negative findings among the pathologically positive findings. A total of 313 patients had at least one LNL with clinical and pathological information.

| HPV+ |  |  |  |  |  |
| --- | --- | --- | --- | --- | --- |
| LNL | Available entries | Path. positives | False neg | Occult mets. rate(%) | 95% CI |
| Total | 789 | 243 | 61 | 24.7% | [19.7, 30.1] |
| I | 111 | 11 | 7 | 63.6% | [35.2, 92.1] |
| II | 186 | 151 | 15 | 9.9% | [5.2, 14.7] |
| III | 184 | 53 | 20 | 37.7% | [24.7, 50.8] |
| IV | 186 | 13 | 7 | 53.8% | [26.7, 80.9] |
| V | 122 | 15 | 12 | 80.0% | [59.8, 100.0] |
| HPV– |  |  |  |  |  |
| LNL | Available Entries | Path. positives | False neg | Occult mets. rate(%) | 95% CI |
| Total | 461 | 119 | 57 | 48.7% | [39.7, 57.8] |
| I | 64 | 11 | 6 | 54.5% | [25.1, 84.0] |
| II | 112 | 58 | 15 | 25.9% | [14.6, 37.1] |
| III | 112 | 34 | 24 | 70.6% | [55.3, 85.9] |
| IV | 111 | 8 | 7 | 87.5% | [64.6, 100] |
| V | 62 | 6 | 5 | 83.3% | [53.4, 100] |

Considering individual LNLs, occult metastasis rates varied substantially across LNLs in both cohorts. In HPV+ patients, the lowest rate was observed in LNL II (9.9% [5.2–14.7]), the most frequently involved level. Higher rates were observed in the less frequently involved levels LNLs III, IV, and V. However, these estimates were associated with wide confidence intervals, reflecting limited numbers of pathologically positive and clinically false-negative cases. In HPV– patients, level-specific occult metastasis rates were higher in most LNLs, particularly in LNLs II–IV.

### 3.5 Occult Metastasis Risk

Occult metastasis risk was evaluated for three ipsilateral clinical diagnoses: clinically negative LNLs II– IV, clinically positive LNL II with clinically negative LNLs III and IV, and clinically positive LNLs II and III with clinically negative LNL IV.

Table 3 summarizes the resulting level-specific occult metastasis risks. Among patients with clinically negative ipsilateral LNLs II–IV, occult risk was highest in LNL II in both cohorts (HPV+: 44.1% [27.4– 60.8]; HPV–: 30.0% [17.3–42.7]). Occult risk was lower in LNL III (HPV+: 14.7% [2.8–26.6]; HPV–: 16.0% [5.8–26.2]) and low in LNL IV (HPV+: 2.9% [0.0–8.6]; HPV–: 4.0% [0.0–9.4]). Among patients with clinically positive LNL II but negative LNLs III and IV, occult risk in LNL III was lower in HPV+ than in HPV– patients (14.7% [7.1–21.4] vs. 34.9% [20.6–49.1]), whereas occult risk in LNL IV remained low in both cohorts (2.2% [0.0–5.2] vs. 7.0% [0.0–14.6]). Among patients with clinically positive LNLs II and III but clinically negative LNL IV, occult risk in LNL IV was similar between HPV+ and HPV– patients (13.8% [1.2–26.3] vs. 14.3% [0.0–32.6]).

**Table 3:** Occult metastasis risk in clinically negative ipsilateral LNLs, stratified by HPV status and clinical involvement diagnosis. The analysis was restricted to patients with both clinical and pathological diagnoses available for ipsilateral LNLs II, III, and IV. FN denotes clinically negative but pathologically positive LNLs, TN denotes clinically and pathologically negative LNLs, and *N* = FN + TN. Risk was calculated as FN/(FN+TN). Values in brackets indicate 95% Wilson confidence intervals.

| Clinical diagnosis | LNL | HPV+ |  |  |  | HPV− |  |  |  |
| --- | --- | --- | --- | --- | --- | --- | --- | --- | --- |
|  |  | FN | TN | N | Risk | FN | TN | N | Risk |
| II–IV negative | II | 15 | 19 | 34 | 44.1% [27.4–60.8] | 15 | 35 | 50 | 30.0% [17.3–42.7] |
|  | III | 5 | 29 | 34 | 14.7% [2.8–26.6] | 8 | 42 | 50 | 16.0% [5.8–26.2] |
|  | IV | 1 | 33 | 34 | 2.9% [0.0–8.6] | 2 | 48 | 50 | 4.0% [0.0–9.4] |
| II positive; III&IV negative | III | 13 | 78 | 91 | 14.3% [7.1–21.4] | 15 | 28 | 43 | 34.9% [20.6–49.1] |
|  | IV | 2 | 89 | 91 | 2.2% [0.0–5.2] | 3 | 40 | 43 | 7.0% [0.0–14.6] |
| II&III positive; IV negative | IV | 4 | 25 | 29 | 13.8% [1.2–26.3] | 2 | 12 | 14 | 14.3% [0.0–32.6] |

The three clinical diagnoses shown in Table 3 represent the most clinically relevant involvement patterns and included 261 of these 313 patients; the remaining patients had other clinical involvement patterns and were not tabulated separately.

## 4 Discussion

In recent years, there has been increasing interest in personalized and volume-de-escalated elective nodal irradiation strategies for OPSCC, including studies of unilateral versus bilateral elective treatment, HPV-associated de-escalation approaches, and model-informed CTV-N personalization [27, 7, 8]. In this context, an important open question is whether HPV+ and HPV– OPSCC differ sufficiently in lymphatic spread or occult metastasis risk to motivate different elective nodal treatment volumes.

### 4.1 Prevalence of lymph node involvement

In this multi-institutional cohort of 780 patients with untreated OPSCC, the most prominent difference between HPV+ and HPV– disease was a higher frequency of ipsilateral LNL II involvement in HPV+ patients, particularly in early T-stages. Also for LNL III, HPV+ patients showed higher involvement for early T-stage, but not for late T-stage. Involvement of LNLs I, IV, V, and VII showed no consistent HPV-related differences after stratification by T-stage. Contralateral involvement was comparable between HPV+ and HPV– patients after stratification by midline extension.

These findings are consistent with previous reports describing more frequent lymph node metastases in HPV-associated disease [9, 17, 18]. Hope et al. [18] recently reported level-specific nodal involvement in an HPV-stratified OPSCC subset of 960 patients from the RADCURE cohort using automated extraction of gross nodal targets from radiotherapy treatment plans. Consistent with our findings, they observed higher ipsilateral LNL II involvement in HPV+ patients, particularly in early T-stages. However, their analysis also showed higher ipsilateral LNL III involvement in HPV+ patients across T-categories, and modestly higher involvement of LNLs IV and V. For contralateral involvement, Hope et al. reported similar overall rates between HPV+ and HPV– patients, although some T-stage-specific differences were observed for individual LNLs. These differences may partly reflect methodological differences, including the assignment of nodes spanning multiple levels to all involved levels, as well as the use of radiotherapy treatment plans to define nodal involvement rather than a level-by-level diagnostic consensus. In addition, HPV– T-stage strata were relatively small, particularly for T1 disease. Thus, while both studies show more frequent ipsilateral LNL II involvement in HPV+ OPSCC, Hope et al. suggest more extensive ipsilateral nodal involvement across additional levels than observed in our cohort.

What our analysis adds to the literature is a more detailed investigation of the conditional probability of the involvement of downstream LNLs given the state of the upstream level, which to our knowledge is not reported in previous studies. The distribution of LNL involvement can be decomposed into the probability of involvement of an upstream LNL and the conditional probabilities of involvement of downstream LNLs. For example, the joint probability of involvement in LNLs II, III, and IV can be written as *P* (*X*_*II*_, *X*_*III*_, *X*_*IV*_) = *P* (*X*_*II*_)*P* (*X*_*III*_ | *X*_*II*_)*P* (*X*_*IV*_ | *X*_*II*_, *X*_*III*_), where *X*_*v*_ represents a binary random variable that indicates the involvement of LNL *v*. Figure 3 suggests that HPV+ and HPV– patients differ in the marginal probability of LNL II involvement, *P* (*X*_*II*_), but probabilities of involvement of downstream levels conditioned on the state of the upstream level are similar. In that sense, higher probability of LNL III involvement for HPV-associated tumors is explained by a higher probability of LNL II involvement, rather than a different pattern of lymphatic spread. Similarly, Figure 2 suggests that contralateral involvement does not depend on HPV when patients are stratified for the main risk factor, midline extension of the primary tumor.

### 4.2 Risk of occult metastases

Compared with the prevalence of clinically or pathologically detected LNL involvement, the risk of occult metastases is more difficult to determine directly from data. It requires information on both the clinical diagnosis and the pathological involvement of individual LNLs after neck dissection. Such data are limited, because pathological information is available only for surgically treated patients and only for dissected levels. Therefore, personalized estimates of occult metastasis risk are limited to the most common clinical diagnoses such as cN0 patients. Moreover, surgical cohorts may be subject to selection bias and may therefore not necessarily be representative of all patients including definitive radiotherapy patients.

Stanford-Moore et al. [22] reported occult metastases in clinically node-negative (cN0) HPV+ OPSCC patients. They found occult involvement in LNL II in 26.9%, LNL III in 8.7%, and LNL IV in 4.0% of patients. The closest comparison in our analysis is the subgroup with clinically negative ipsilateral LNLs II–IV, in which occult metastasis risk in HPV+ patients was 44.1% [27.4–60.8] for LNL II, 14.7% [2.8–26.6] for LNL III, and 2.9% [0.0–8.6] for LNL IV. Thus, our estimates were higher for LNLs II and III, but comparable for LNL IV. Direct comparison is limited by differences in cohort selection, clinical definitions, and the limited number of patients with paired clinical and pathological information in the relevant subgroup. Moreover, the cohort in the study of Stanford-Moore et al. was dominated by T1-2 (“early”) stages, which could also explain the overall lower involvement rates observed.

The data in Table 3 suggests that, when LNLs II–IV were clinically negative, occult metastasis risks were similar between HPV+ and HPV– patients. Among patients with clinically positive LNL II and clinically negative LNLs III and IV, occult involvement risk in LNLs III and IV was lower in HPV+ patients. Finally, when LNLs II and III were clinically positive, occult risk in clinically negative LNL IV was similar between HPV+ and HPV– patients. Hence, the observed differences in occult metastasis risks between HPV+ and HPV– tumors were not consistent. In addition, the estimates are associated with large uncertainties, and the data is not stratified according to T-stage. Hence, the findings must be interpreted with caution.

The discrepancy between clinical and pathological involvement has been described in the literature via the sensitivity and specificity of the diagnostic modalities. In our data, a smaller proportion of pathologically involved LNLs was clinically (false) negative in HPV+ tumors, i.e. the sensitivity for clinically detecting lymph node metastases appears higher in HPV+ patients compared to HPV– patients (Table 2). However, this observation must also be interpreted carefully. The lower false-negative rate in HPV+ patients is in part driven by the high prevalence of clinical involvement in LNL II, which may in many cases trigger the patient’s diagnosis of OPSCC, rather than the primary tumor itself. The observation that the proportion of occult metastases increases gradually from LNL II to LNL III and IV shows general limitations of the concept of sensitivity in this context as previously discussed.

### 4.3 Relation of occult metastasis risk and prevalence of involvement

Observed LNL involvement and occult metastasis risk are related but distinct quantities. The risk of occult metastases can be understood as the probability of involvement given that the LNL is clinically negative. This probability depends on two components: the underlying probability of LNL involvement and the sensitivity and specificity of clinically detecting metastases. In previous work, these two aspects have been combined in statistical models to estimate the risk of occult metastases [26]. As a typical example, one may consider a patient with clinical involvement of LNL II, confirmed by fine needle aspiration, and a given T-stage and midline extension status. In such a patient, the risk of occult disease in clinically negative downstream LNLs depends both on the conditional probability of downstream involvement and on the ability of the clinical diagnostic work-up to detect existing disease. The data presented in Figure 3 suggest that conditional probabilities of downstream involvement, given the state of the upstream LNL, were similar between HPV+ and HPV– patients. Considering this aspect, one would not expect a higher probability of occult involvement in downstream LNLs in HPV+ disease once the clinical involvement pattern is taken into account. At the same time, the occult metastasis rate among pathologically positive LNLs was lower in HPV+ than in HPV– patients (Table 2), i.e. the sensitivity of clinically detecting lymph node metastases appears higher for HPV+ tumors. Considering this aspect, one would expect a lower probability of occult metastases for HPV+ tumors.

## 5 Conclusions

In this large, multi-institutional cohort, HPV+ OPSCC patients more frequently presented with ipsilateral LNL II involvement, particularly in early T-stage disease. However, conditional probabilities of downstream involvement in LNLs III and IV, conditioned on the involvement of the upstream LNL, were similar between HPV+ and HPV– patients. Similarly, contralateral involvement was similar between HPV+ and HPV– patients after stratification by midline extension. In that sense, HPV+ and HPV– patients did not show different pattern of lymphatic spread.

The analysis of occult metastases suggested a higher sensitivity of clinically detecting lymph node metastases in HPV+ patients, i.e. a lower fraction of pathologically positive LNLs was clinically undetected. However, direct estimates of the risk of occult disease for the most common clinical involvement patterns were not uniformly lower in HPV+ patients and were associated with large uncertainties due to the limited availability of pathological data. Taken together, the data on occult metastases has too many limitations to draw definitive conclusions. However, the present data does not provide evidence that elective nodal treatment volumes should differ between HPV+ and HPV– OPSCC.

## Data Availability

all data are available on lyprox.org

https://lyprox.org

## 6 CRediT authorship contribution statement

**Yoel Pérez Haas**: Writing – original draft, Data curation Visualization, Project administration, Investigation, Formal analysis. **Roman Ludwig**: Writing – review & editing, Data curation. **Esmée L. Looman**: Writing – review & editing. **Panagiotis Balermpas**: Writing – review & editing, Supervision. **Sergi Benavente**: Writing – review & editing, Data curation. **Adrian Schubert**: Writing – review & editing, Data curation. **Dorothea Barbatei**: Writing – review & editing, Data curation. **Laurence Bauwens**: Writing – review & editing, Data curation. **Olgun Elicin**: Writing – review & editing, Data curation. **Matthias S. Dettmer**: Writing – review & editing, Data curation. **Roland Giger**: Writing – review & editing, Data curation. **Vincent Grégoire**: Writing – review & editing, Data curation. **Jan Unkelbach**: Writing – original draft & editing, Supervision, Project administration, Funding acquisition, Conceptualization, Resources.

## 7 Acknowledgements

This work was supported by the clinical research priority program (CRPP) “Artificial intelligence in oncological imaging” of the University of Zurich, by the Swiss cancer research foundation under grant KFS 5645–08–2022, and by the Swiss National Science Foundation under grant CR00-5-239827.

## Notes

### Competing Interest Statement

The authors have declared no competing interest.

### Author Declarations

All the data has been published before and is available on lyprox.org

